# COVID-19 Scenarios: an interactive tool to explore the spread and associated morbidity and mortality of SARS-CoV-2

**DOI:** 10.1101/2020.05.05.20091363

**Authors:** Nicholas B Noll, Ivan Aksamentov, Valentin Druelle, Abrie Badenhorst, Bruno Ronzani, Gavin Jefferies, Jan Albert, Richard A Neher

**Affiliations:** Biozentrum, University of Basel; Swiss Institute of Bioinformatics, Basel, Switzerland; unaffiliated contributors; Department of Clinical Microbiology, Karolinska University Hospital, Stockholm, Sweden; Department of Microbiology, Tumor and Cell Biology, Karolinska Institutet, Stockholm, Sweden

## Abstract

The ongoing SARS-CoV-2 pandemic has caused large outbreaks around the world and every heavily affected community has experienced a substantial strain on the health care system and a high death toll. Communities therefore have to monitor the incidence of COVID-19 carefully and attempt to project the demand for health care. To enable such projections, we have developed an interactive web application that simulates an age-structured SEIR model with separate compartments for severely and critically ill patients. The tool allows the users to modify most parameters of the model, including age specific assumptions on severity. Infection control and mitigation measures that reduce transmission can be specified, as well as age-group specific isolation.

The simulation of the model runs entirely on the client side in the browser; all parameter settings and results of the simulation can be exported for further downstream analysis. The tool is available at covid19-scenarios.org and the source code at github.com/neherlab/covid19_scenarios.

---

The novel coronavirus SARS-CoV-2 was first detected in the city of Wuhan within the Hubei province of China at the end of December 2019 (Li *et al*., 2020). In the following months, SARS-CoV-2 has shown to be highly transmissible - the basic reproductive number, *R*_0_, has been estimated to be within 2-3 (Riou and Althaus, 2020; Zhang *et al*., 2020) with an estimated serial interval of 5-7 days (Ganyani *et al*., 2020; Nishiura *et al*., 2020). The basic reproduction number likely varies between communities and is affected by intervention measures. The illness caused by SARS-CoV-2 infection, COVID-19, clinically presents with a large variance of symptoms that range from mild and asymptomatic infection to acute severe respiratory illness. The clinical presentation of the infection strongly depends upon patient age (Surveillances, 2020) and certain comorbidities (Fang *et al*., 2020). The WHO declared the COVID-19 outbreak a pandemic on March 11th, 2020 (The WHO COVID-19 group, 2020). As of April 20th, 2020, there have been over 2.4 million confirmed COVID-19 cases from 210 countries.

A critical component of the global response to the COVID-19 pandemic is the possibility to explore different scenarios for local outbreaks within communities across the world using mathematical modelling. Modelling is important not only to guide governmental public health policy but also to inform hospital readiness and educate the general public on the importance of social distancing efforts. The spectrum of models used to analyze COVID-19 outbreaks ranges from computationally intensive agent-based simulation (Neil M Ferguson, 2020), variants of SIR/SEIR models (Kermack *et al*., 1927), to phenomenological curve fitting approaches (“IMHE COVID-19 forecasting team” and Murray, 2020). However, traditional epidemiological modelling protocols do not scale for a global pandemic - modelling has to be done on a region-by-region basis. Thus, to make such modeling widely available, we have developed an interactive, online tool that allows users to efficiently explore COVID-19 scenarios based upon different epidemiological assumptions and potential mitigation strategies. The dynamics are modelled by an age-stratified SEIR model, with additional novel compartments that correspond to hospital and ICU utilization with finite capacity.

Our deterministic approach strikes a compromise between the accuracy of the approximation of the outbreak dynamics and the speed of the simulation. On a typical modern computer and browser, the simulation will complete in under one second such that many different parameter values can be explored interactively. The output of the model is a time series of simulated COVID-19 infections, hospitalizations, and ICU usage. Surveillance data such as case counts, COVID-19-related fatalities, and hospitalizations can be compared to the model output when such data are available. Additionally, we utilize these data to estimate a few basic parameters for each provided scenario to provide reasonable starting points for further parameter explorations. However, we stress that the focus of this tool is on the exploration of scenarios and not on parameter inference.

We have designed our tool with the following principles: (i) users should be able to interact dynamically with the simulation such that changing underlying assumptions manifests instantly in the results, (ii) empirical surveillance data should be plotted with the simulation results to allow for easy assessment of parameter assumptions, (iii) results should be easily shareable via URLs, exported raw data, and parameter files. Our tool, COVID-19 Scenarios, was first released on March 9, 2020 and was one of the first publicly available interactive models. It has been utilized consistently throughout the COVID-19 pandemic, averaging roughly 8 thousand page loads per day. Since we first released, we have been dedicated to improving the tool, in both its underlying scientific accuracy as new data emerged, as well as the overall user experience. All source code and the aggregated surveillance data are made freely available through GitHub.

## BASIC MODEL

We approximate the dynamics of a COVID-19 outbreak using a generalized SEIR model in which the population is partitioned into age-stratified compartments of: susceptible (S), exposed (E), infected (I), hospitalized (H), critical (C), ICU overflow (O), dead (D) and recovered (R) individuals (Kermack *et al*., 1927). The progression of illness is approximated by the following compartment transitions: susceptible individuals are exposed to the virus by contact with an infected individual; exposed individuals progress towards a infectious state; infectious individuals either recover without hospitalization or progress towards a severe illness that requires hospitalization; hospitalized individuals either recover or worsen towards a critical state; individuals with a critical illness either transition to the ICU or, if the hospital is at capacity, to an “overflow” compartment and either return to the hospital state or die; recovered individuals can not be infected again. See Fig. 1 for an illustration of the model. We note that direct comparisons between the model predictions and available surveillance data are difficult since only a fraction of cases are confirmed by a positive test and this fraction various between regions. The number of COVID-19 deaths is often a more robust measure.

**Figure 1.**
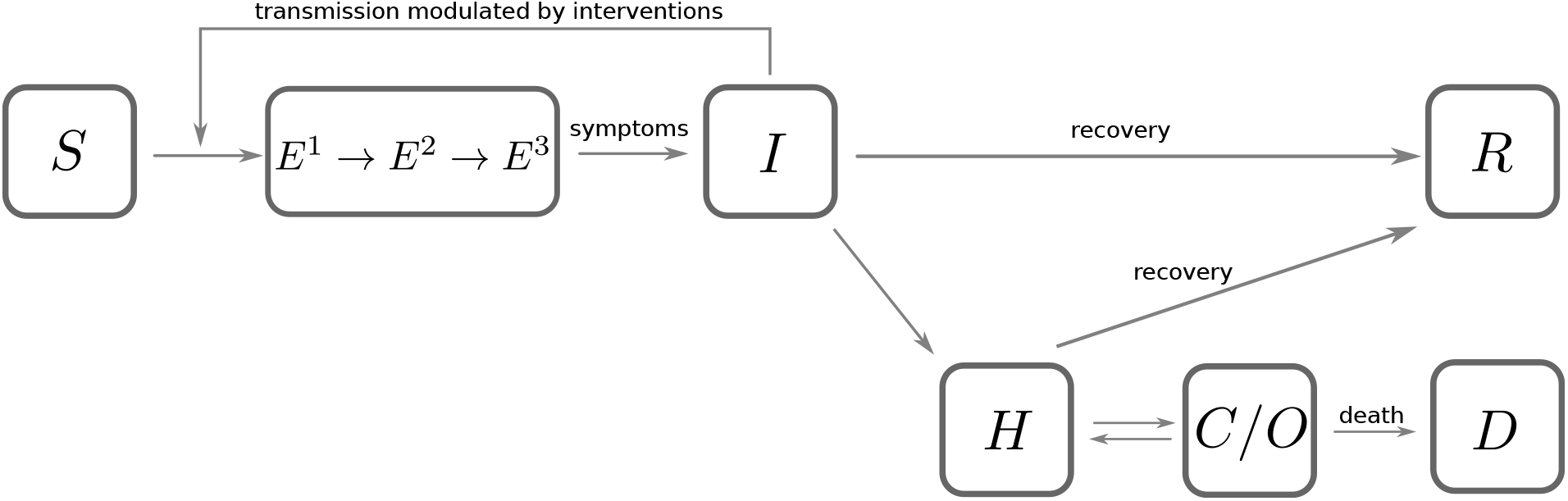
Diagram of model. A schematic illustration of the underlying model and allowed transitions. *S*, *E^i^*, *I*, *R*, *H, C, O* and *D* represent the susceptible, exposed, infectious, recovered, hospitalized, critical, overflow, and fatal compartments of the model. Each compartment is further stratified by age demographics.

Let *a,b* ∈ [1, 2,…, *N_a_*] denote the different age classes of each compartment. The parameters of the model fall into three broad categories: a time-dependent infection rate *β_a_(t);* the rate of transition out of the exposed, infectious, hospitalized, and critical/overflow compartments *γ_e_*, *γ_i_*, *γ_h_*, and *γ_c_* respectively; and the age-specific fractions *m_a_, c_a_* and *f_a_* of mild, critical, and fatal infections respectively. Below, we expound upon each class of parameter.

The rate of transmission, *β_a_(t)*, is nominally determined by both the basic reproductive number *R*_0_ and the time period of patient infectivity 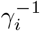. Additionally, the rate of transmission can be effectively slowed by mitigation efforts (e.g. social distancing), which we account for phenomenologically by a multiplicative factor 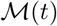 (see below). Lastly, empirical data shows a strong, consistent seasonal variation of the four endemic coronaviruses suggesting similar seasonality in the transmissibility of SARS-CoV-2 (Neher *et al*., 2020) Taken together, the rate of transmission is modelled by

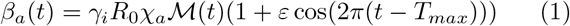

where *χ_a_* models specific demographic isolation, and *ε* and *T_max_* denote the (currently unknown) amplitude of seasonal variation in transmissibility and the time of the year of peak transmission respectively.

After an individual is infected (i.e. was exposed) it takes some time before the individual itself is infectious. In our model, the average value of this latency is given by 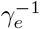. The incubation time of COVID-19 has been estimated to be well approximated by an Erlang distribution (Lauer *et al*., 2020). As such, we approximate the distribution of incubation times within our framework by chaining three exposed states in which the mean time to pass through all three states is 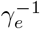. The mean infectious time of a COVID-19 case is 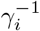 which together with the incubation period 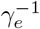 defines the serial interval. The residence times in the remaining compartments are assumed to be exponentially distributed and thus taken to be a singular state.

As noted above, the fraction of COVID-19 infections that are asymptomatic/mild, severe cases which progress to a critical state, and critical cases that are fatal are denoted as *m_a_, c_a_*, and *f_a_*, respectively (see below for more detail). However, it is important to consider the effects of hospital capacity and overutilization in forecasting potential scenarios. Finite hospital resources and staffing acutely impact the outcome for critical COVID-19 patients and thus the overall COVID-19-related fatalities. We phenomenologically capture this effect by introducing non-linear constraint of a finite number of ICU beds C that can accommodate critical patients. Once the number of critical cases exceeds this parameters, additional critical cases are redirected to an “overflow” compartment. We take the mortality rate of an overflow patient relative to a patient with an ICU bed to be ξ.

With all parameters explicitly defined, our full model can be written

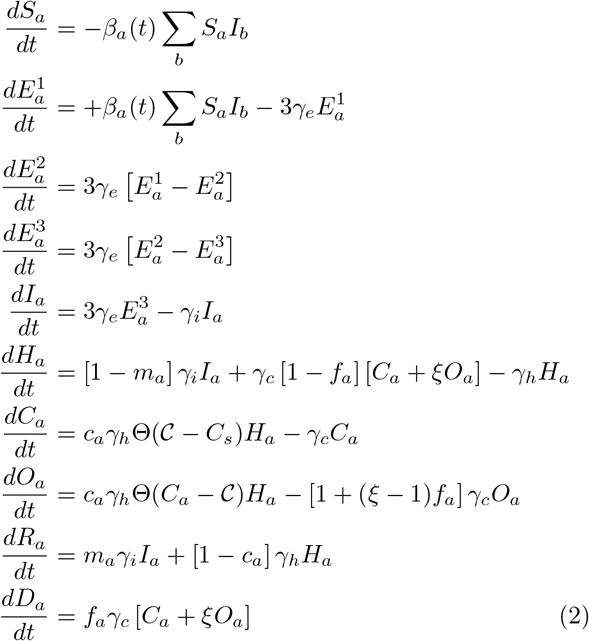

where Θ(*x*) denotes the Heaviside step function and imposes the finite constraint of hospital resources.

The model currently does not allow for reinfection. Evidence from other coronaviruses suggests that infection elicits immunity that lasts for at least a year (Callow *et al*., 1990). Whether this also holds for SARS-CoV-2 is not yet clear, but reinfection and herd immunity are of minor relevance in the early phase of a pandemic (Neher *et al*., 2020). Reinfection might be added to the model in the future if evidence accumulates that it is important.

## USER INTERFACE AND ADJUSTABLE PARAMETERS

Epidemiological models, including the one defined by Eqn 2, have dozens of parameters, many of which are not accurately known and difficult to measure. Additionally, each model dramatically simplifies reality; our parameters are phenomenological summaries of the “true” heterogeneous dynamics. Therefore, we give the user control over *all* model parameters in order to facilitate the exploration of the dependence of the predicted results on the input parameter values, see Fig. 2A for a screenshot of the UI. We note that users specify timescales instead of rates in the UI, e.g. 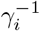 corresponds to the “Infectious period” input box of Fig. 2A, as we felt timescales are easier to directly interpret. For ease of use, the web application has presets for many countries and states that can be used as a *starting point* for exploration.

**Figure 2.**
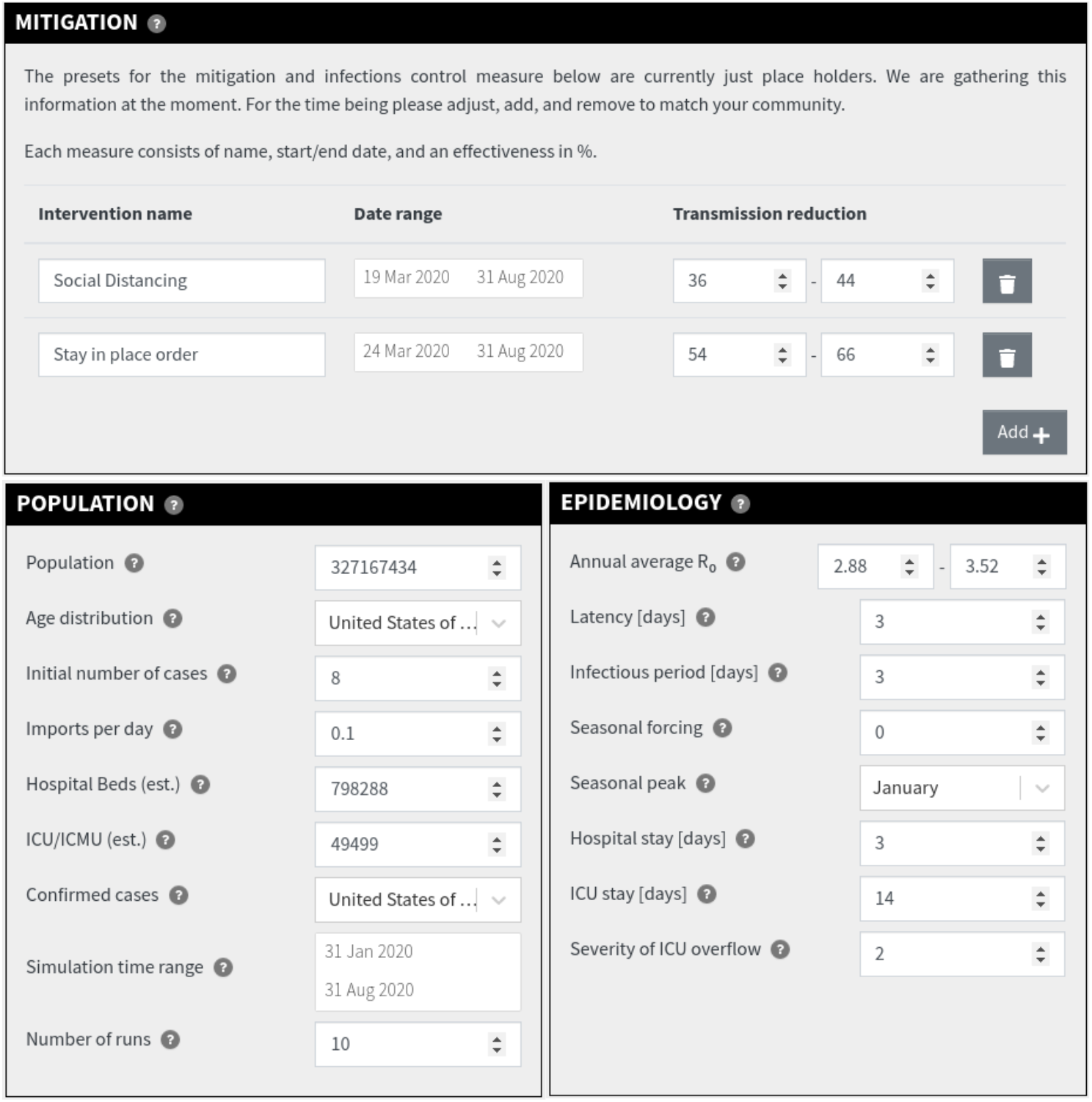
User interface for model parameters and mitigations. A) An example screenshot of the UI of the web application used to vary model parameters. On the left, we have grouped parameters of the population under study, e.g. the initial number of cases and the size of population. Conversely, on the right, we grouped phenomenological parameters related to COVID-19 epidemiology. All numbers can be entered manually with a keyboard or stepped with the scrollbox. B) Individual mitigation measures can be added or removed from the model via the the shown interface. Each intervention has a unique name, a range of times it is applied for, and a range of possible efficacies. The net mitigation on COVID-19 transmission is calculated via Eq. 3

### Interventions

In order to model both the historical transmission of COVID-19 and project its further spread, one must model the enacted social distancing measures, case isolation, and quarantine policies. As such, our model gives the user the ability to specify individual interventions, indexed by *α*, with a well-defined start and end date and an “effectiveness” *∊_α_* ∈ [0,1] parameter that quantifies the mitigation’s multiplicative effect on rate of transmission. See Fig. 2B for an depiction of the UI for the input of different mitigation measures. At each point in time, the cumulative efficacy of all interventions is calculated as

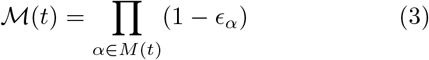

where the product runs over all measures 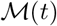 in effect at time t. In the absence of mitigation strategy, 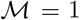1. The overall mitigation efficacy modulates COVID-19 transmissibility as seen in Eqn. 1.

Additionally, our model allows for the input of simple, time-independent age-specific isolation measures. As can be seen in Fig. 3, we provide a column for “age-specific” isolation. These numbers result in a reduction in exposure of individuals from specific age-groups to the general population. For example, this feature could explore the effect of measures specific for the elderly.

**Figure 3.**
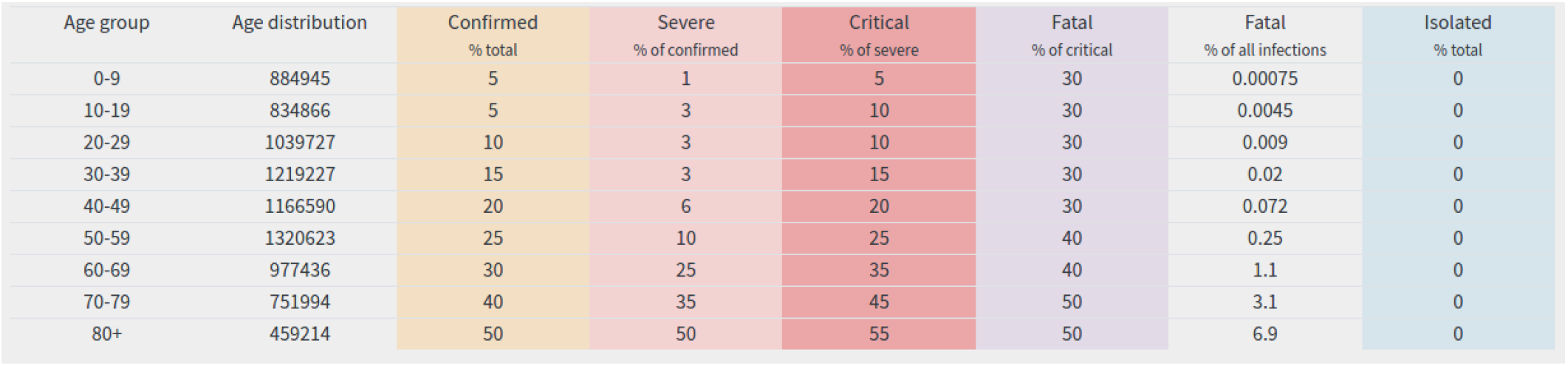
Age-dependent parameters. Parameters that depend on patient age are summarized in a table that contains the distribution of age groups, severity parameters, and age-specific isolation. The severity parameters are approximately based on data by Surveillances (2020). The first column “confirmed” is our assumption on what fraction of total infections in the different age groups enter as cases in the data analyzed in Surveillances (2020). Young individuals are often asymptomatic and hence less likely to be tested. The following columns specify what fraction of confirmed cases fall severely ill and require medical attention, what fraction of the former fall critically ill and requires intensive care, and lastly hat fraction of critically ill patients die. The implied infection fatality rate is given in the second to last column.

### Severity parameters and demographics

The clinical outcome of a COVID-19 infection strongly depends on the age of the patient (Zhou *et al*., 2020). Hence, the overall burden of a COVID-19 epidemic within a given region strongly depends on the age demographics of the population. In order to facilitate the integration of such effects within the model, we aggregated age distributions for most countries, obtained from the UNSD database API (United Nations Statistics Division, 2020) with a custom python script, to provide as presets. Additionally, we allow for custom age distributions to be specified within the UI, see Fig. 3. The provided age distributions determine the *fraction* of people in each age group in the simulation.

The Chinese CDC provided extensive statistics of severity of COVID-19 in different age groups (The Novel Coronavirus Pneumonia Emergency Response Epidemiology Team, 2020), broadly compatible with estimates by (Verity *et al*., 2020). We used these data to parameterize the expected burden on health care systems. Our severity assumptions are summarized in a editable table in the tool, shown in Fig. 3. Each column can be edited and changed if users want and the implied infection fatality for each group is calculated. Elements of the table directly correspond to model parameters: (i) *m_a_* is the product of the percentage confirmed and the complement of the severe fraction, (ii) *c_a_* is set by the critical column, and (iii) *f_a_* is set by the fatal fraction.

### Parameter uncertainty

The dynamics of an exponentially growing process such as the COVID-19 pandemic is naturally most sensitive to the growth rate. In the context of the model, the growth rate of infections is primarily a function of the basic reproductive number *R*_0_ and the societal interventions enacted to slow the spread of COVID-19. Additionally, it is *a priori* difficult to know the efficacy of mitigation measures. COVID-19 scenarios therefore allows the user to specify *ranges* for *R*_0_ as well as for the efficacy *∊_α_* of mitigation measures, see Fig. 2AB for an example of each. The tool will randomly sample a user-specified number of parameter combinations uniformly from these ranges (by default set to 10 combinations). The outputted results display the median as well as a shaded area denoting the 20th and 80th percentile, see Fig. 4 for an example of the displayed results.

**Figure 4.**
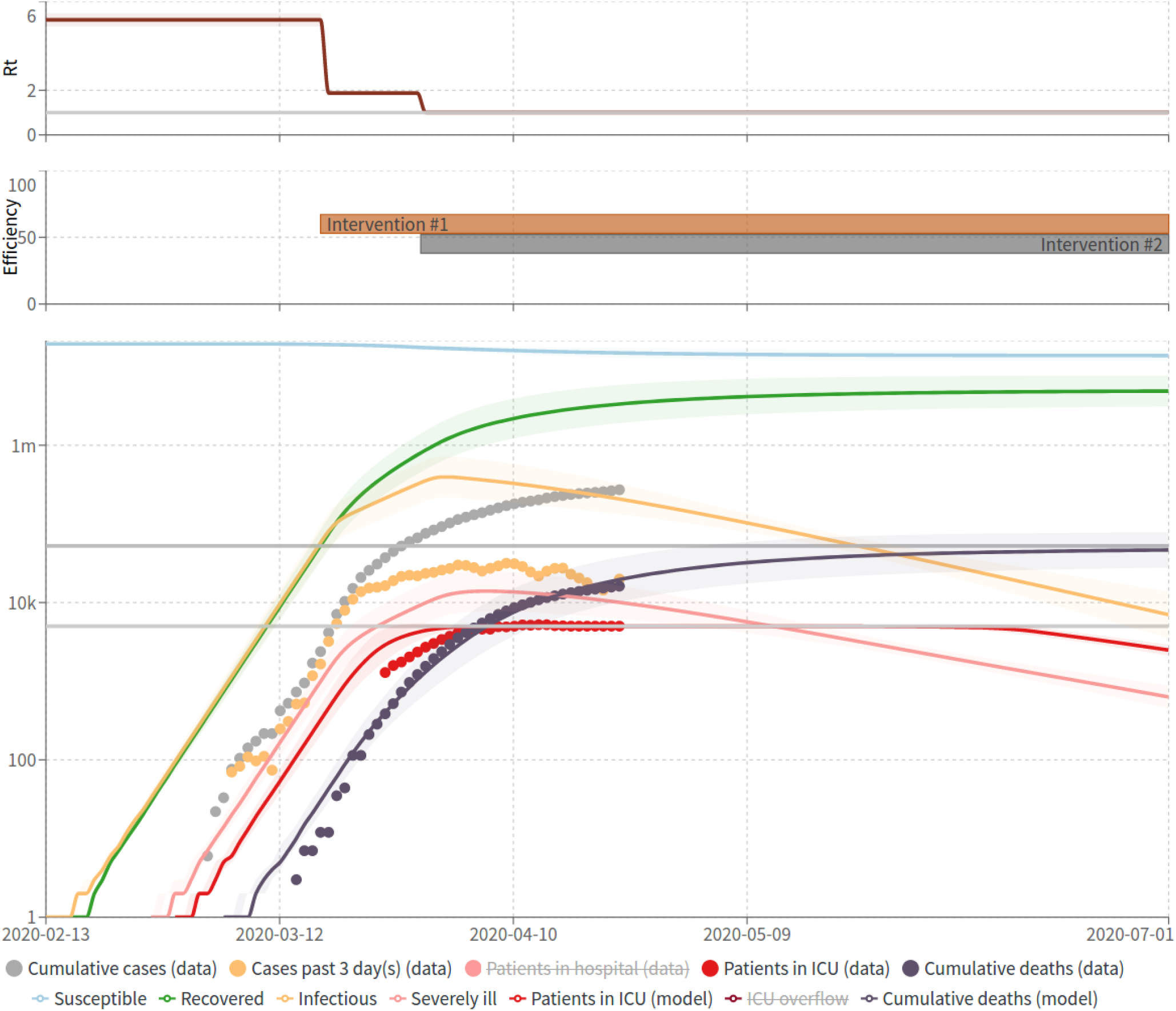
Example simulation results for New York City. (top) A plot of the time-dependent effective reproductive number. This is controlled via mitigation interventions and seasonality. The colored line corresponds to the median and the shaded area is bounded by the 20% and 80% percentiles. (middle) Plot that shows both the interval (length) and ranges of possible efficacies (width) for the applied mitigation interventions. (bottom) Plot that shows the resulting trajectories of all compartments. The colored line for each compartment shows the median trajectory while the shaded area is bounded by the 20% and 80% percentiles. Additionally, the aggregated case count data is plotted (where available) as individual points. The display of individual compartments can be toggled by clicking on the legend.

### Model results

The primary result of the tool are trajectories of the number of cases, people in need of hospitalization, and fatalities, see Fig. 4 for an example of the predictions for New York City. Additionally, all predicted trajectories can be exported as a single age-stratified table for further downstream analysis. A short executive summary of the results can additionally be printed to PDF. Where available, the app graphs the recent cases counts, deaths, and hospitalizations for the community under study on the plot with the model results. The surveillance data enables the user to adapt parameters to tune the simulation to the data, see Fig. 4. Once the model fits past data, the user can explore future scenarios by adjusting interventions and seasonality.

## ESTIMATION OF PARAMETERS FROM DATA

While COVID-19 Scenarios is not intended as an inference tool, we nevertheless provide parameter presets that are estimated from empirical data. The primary intent behind fitting to data is not to provide values with high confidence, but rather to facilitate the immediate utility of our tool for different scenarios from across the globe with reasonable presets. We note that care must be taken to not *overfit* the data; there are many more parameters of the model than features within the available data. Furthermore, the testing and reporting patterns are heterogeneous across regions, as well as change over time which ultimately distort the raw numbers.

We therefore elect to only estimate three model parameters for each region: (i) *R_0_*, not solely a property of the virus but also the social structure of the population, (ii) the initial date of the epidemic *t_min_*, and (iii) the size of the initial cluster *I*_0_. In addition, we preset mitigation measures that set in when case-counts rise above certain levels. Again, these are not meant as fit parameters but as templates to be adjusted by the user. We assume the remaining parameters don’t vary across regions. These have to be adjusted by the user if the data or other information suggests values different from the defaults.

We try to fit data solely from the onset of the epidemic prior to mitigation efforts from individual regions. Due to the heterogeneity of both the timing and efficacy of policies implemented across the regions provided for interactive exploration, we opted for a simple solution. As more data from more regions become available, we might fit more parameters to observations.

## EMPIRICAL DATA

Both the initial estimates for scenario values, as well as the interactive calibration of the model require empirical observations of COVID-19 infections and hospitalizations. Due to the scope of scenarios provided, we utilize a number of online resources to aggregate information on new COVID-19 cases, deaths, and hospitalizations. These resources include the daily updated case counts by ECDC (European Centre for Disease Control, 2020), the US COVID tracking project (The COVID tracking project, 2020), other official governmental agencies from around the world, and data aggregated by volunteers. A full list of all sources we use can by found in the file data/sources.json in https://github.com/neherlab/covid19_scenarios. The case empirical data in the app is updated every 2-3 days.

## IMPLEMENTATION AND AVAILABILITY

COVID-19 Scenarios is implemented as a single-page web application using React web framework, Typescript and numerous packages from Node.js ecosystem. The simulation itself runs on the client side in a WebWorker, to ensure interactivity during the computation. The application can be hosted on any static web-server, or run locally. We host the latest release version publicly on AWS infrastructure, accessible at https://covid19-scenarios.org.

Data fetching, processing, parameter estimation and scenario generation is implemented using Python and common data science packages, as an additional build step. The full source code is available under MIT license on GitHub at https://github.com/neherlab/covid19_scenarios. Instructions on how to run the application are documented there. COVID-19 Scenarios was first released on March 9 2020 and has been updated consistently since.

## DISCUSSION

Countries, states, and communities across the world have to plan and prepare for the outbreaks and potential reemergence of COVID-19. Many countries have expert research groups that develop tailored models and sophisticated inferences (Kucharski *et al*., 2020; Neil M Ferguson, 2020) to predict individualized outcomes. However, governments and other public organizations without such availability need a flexible tool that models local outbreaks, explore the effect of interventions, and can compare results to past dynamics in an interactive workflow. This is the gap that COVID-19 Scenarios has and continues to fill. To date, we average roughly 8 thousand page loads per day (we don’t track users, but estimated these numbers of from CloudFront usage statistics and the number of requests per page load). These requests come from more than 50 countries, with most visitors coming from the USA, Germany, Switzerland, Russia, Austria, and the UK.

In order to estimate the potential future burden on the health care system, users need flexible ways to adjust demographic parameters in addition to local public health care policies (who gets admitted to the ICU, how long are patients hospitalized). At the same time, sensible defaults are required to provide a useful starting point for exploration. COVID-19 Scenarios was written with the explicit purpose to aid in this regard.

The past few months have shown that social distancing measures can effectively slow the spread of COVID-19(Wang *et al*., 2020). The future trajectory of COVID-19 will therefore primarily depend upon the level of social distancing and infection control that is maintained. COVID-19 Scenarios therefore cannot confidently *predict* outcomes, but rather help to explore *potential* future scenarios under specific assumptions made by the user. This difficulty is further compounded by the fact that the prediction of absolute numbers are exceedingly sensitive to small variations in input parameters. Due to nature of exponential growth experienced within an epidemic, a small uncertainty in either the growth rate or initial date will naturally result in large uncertainty in case numbers. Therefore, it is critical that these uncertainties are communicated effectively to policy makers. We therefore allow the user to specify plausible ranges for the parameter Ro and the efficacy of the interventions. In case of several interventions, this results in a high dimensional space of possibilities that we sample uniformly. Percentiles of the sampled results are displayed to capture the range of potential outcomes.

We stress that in addition to parameter uncertainty, a simple SEIR model is a drastic abstraction and simplification that does not capture the full complexity and heterogeneity of the outbreak. Nevertheless we hope that the tool is helpful for understanding the dynamic of the outbreak and exploring the effect of past and future interventions.

## Data Availability

All source code and data are available on github.com/neherlab/covid19-scenarios

## Acknowledgement

We gratefully acknowledge input from members of the lab, Adam Kucharski, Rosalind Eggo, and Christian Althaus. Nils Ole Tippenhauer and Aitana Lebrand have helped to parse, aggregate, and update surveillance data from various sources. In addition, we received many invaluable contributions from the open source community. vercel.com has supported the development with free access to their tool-stack.

